# Sample pooling, a population screening strategy for SARS-CoV2 to prevent future outbreak and mitigate the “second-wave” of infection of the virus

**DOI:** 10.1101/2020.10.29.20204974

**Authors:** S. Sawarkar, A. Victor, M. Viotti, S. Haran, S. Verma, D. Griffin, J. Sams

**Affiliations:** Rosa Scientific, Princeton, NJ, USA; Zouves Foundation for Reproductive Medicine, San Francisco, CA, USA; University of Kent, Canterbury, Kent, UK

## Abstract

**Problem:** How do we manage treatment and stabilization in clinical settings and static population communities like assisted living facility settings of their patient or resident populations during and post the SARS-CoV-2 pandemic?

**Scope:** This proposal explores the possible and predicted changes to standard operating procedures to the facility management and associated landscape and focuses on series of deployments, during and post peak SARS-CoV-2 activity, and will outline possible models for the current medical facility model that we operate with. This article primarily focuses on non-emergency facility management.

**Assumptions and understanding of the field:** With a reduction in the numbers nationally, patients are highly motivated and likely to seek non-emergency and planned medical procedural treatment as early as possible as social distancing measures are eased and restrictions on non-urgent procedures are lifted.

**Conclusions and next steps:** An initial pan-national shutdown and suspension of services was necessary in an effort to ensure that essential medical services and resources were not strained. The authors feel that a strategic resumption of regular non-emergency treatments around the United States and continued provision of services at care facilities is possible with innovative testing strategies like pooled screening of large populations at a manageable price point. Moreover, pooling as a strategy when used widely, would be extremely effective at predicting outbreaks of the virus and as an effect help in mitigating the spread of the virus in its “second-wave”. We have developed one such innovative pooling strategy that can be easily deployed across laboratories and reduce the cost of population wide COVID-19 testing significantly

## 1. Introduction

SARS-CoV2 is the virus that causes COVID-19. This novel infectious agent has followed a completely unique trajectory of infection and virulence. SARS-CoV-2’s severity has manifested itself in a range of possible symptoms with varying gravities (from mild to fatal), and due to high degree of virulence has been defined as a pandemic by the WHO (WHO, 2020). The early studies out of China have shown a specific at-risk population (median age of death being 75, (Wang, Tang, & Wei, 2020)). The studies have also characterized the treatment options currently deployed as symptomatic treatments only (Sun, Lu, Xu, Sun, & Pan, 2020). The best strategy to contain the spread is to test as much as possible, this however comes at an extremely high cost.

There has not been a major infectious pandemic in a very long a time (apart from the influenza virus pandemic in the year 1918). The last known cases of epidemic-level viral agents that have passed through populations are Spanish Flu and HIV/Aids. Transcontinental migration patterns have exponentially increased and trade and travel is no longer restricted by water boundaries. This has changed the way health systems function during and post pandemic situations. Seasonal influenza is an ineffective model candidate to mimic the COVID-19 response. Influenza is also one of the most well characterized viral agents, as the field has demanded yearly reviews over the last decade.

Health systems and pharmacies have adapted to the yearly revenue model that Influenza provides, and this epidemic is well prepared for every year. Preventive methods such as work-place flu shots, major pharmacies administering flu shots, as well as knowledge and community outreach events have made influenza a well-managed epidemic. For example, proposals for drive through medicine for influenza were documented as early as 2010 for quick, limited spread testing (Weiss, Ngo, Gilbert, & Quinn, 2010).

Most non-emergency groups have initially reacted to COVID-19 by ceasing operations completely. While the authors believe that this was necessary to avoid strain on scarce medical sources, in the more recent weeks, many COVID-19 hotspots have already passed their projected peak requirement of resources. To that end, resumption of regular services should be considered. Moreover, this resumption can be specified based on statewide and local COVID-19 statistics. It is important to note that potential “subsequent waves” might force the providers to put a temporary halt on providing their services.

A study from the University of Oxford, Department of Zoology, mapped the model of infection for the SARS-CoV-2 in the UK population (Lourenco et al., 2020). According to their proposed model, epidemiologically there are three phases of the infection cycle:

1. Initial infections (often undetected, due to the lack of test availability)
2. Rapid growth of infections and increased deaths
3. Slowing down of infections due to the lack of susceptible individuals

This is crucial in understanding where this proposal is most effective. The models proposed by the authors show this triphasic window - which include social distancing, bans and governmental regulations on population movement to be <u>2-3 months</u> (Lourenco et al., 2020). This 2-3 month window of opportunity should involve two major phases of resource deployment:

### A) “Stay at home” Outreach +Telemedicine

Majority providers in the US have rapidly adopted telemedicine platforms and have been performing consults throughout the pandemic. While formal numbers are not available, estimates suggest an overall increase of anywhere between 500-4000% increase in the number of tele-health consults at clinics. Electronic and telephonic outreach has also been adopted rapidly to maintain patient engagement through the lack of in-person visits.

### B) “Post-Quarantine” Proactive Campaign

As peak number of cases decreases, proactive measures to return back to established practice are crucial. Pooling of samples to reduce the cost burden on the testing mechanism has been historically used to prevent outbreaks of diseases viz. Malaria. Here we suggest an innovative approach of a pooling strategy for COVID-19 testing.

To be able to resume services, where suspended, and to be able to keep providing services, its utmost important to be able to identify any asymptomatic carriers in the staff or the patients and visitors. The sooner an asymptomatic or presymptomatic carrier is identified, the better it is for the containment of the spread of the infection, since primary contacts could be traced and quarantined to break the chain. WHO and several international bodies have time and again reiterated the need for wide scale screening. There has been a worldwide push to increase the testing capacity, however these efforts hit a bottleneck in terms of ability of the testing facilities to test number of samples to be tested per day. The prevalent gold standard of the COVID-19 screening is, real-time PCR-based assay. This assay is very sensitive and can detect as few as a single copy of SARS-CoV2 RNA in a microliter of the total RNA isolated from the patient’s throat swab (Product data sheet of the Real time kit TaqPath™ COVID-19 Combo Kit). However, the real-time assays are time consuming and require highly trained scientists to run them assay and interpret the data, which adds up to the high cost and low turnaround time. Any disease that has required screening of a large population has experienced (faced) this bottleneck. About 80 years ago a pooled testing strategy was proposed, resulting in the so-called “Dorfman testing” method. Per this strategy, if the pool (a combined mix of several samples) tests negative, it means that all the samples in the pool are negative. However, even if a single sample in the pool is positive, it results in the whole pool being positive; To further confirm the identity of the positive sample(s) in the pool, each sample needs to be tested individually. Sample pooling or grouping strategies significantly reduce the turnaround time as well as the cost. Since its inception, sample pooling has been used to screen populations for wide variety of diseases in various settings like, in medical clinics, for chlamydia and gonorrhea, influenza and in the field in mosquitoes for the West Nile virus (Gaydos *et al*, 2005; Hourfar *et al*, 2007 and White *et al*, 2001). Here, we present data in support of sample pooling as a financially viable strategy for screening employees and visitors at businesses. This will enable them to reopen post-pandemic and avert any future outbreaks.

## 2. Results

### 2.1 Limit of detection for SARS-CoV-2 RT-PCR Kit

Several countries have been using pool testing as a successful strategy for the population wide screen of SARS-CoV-2 infection (Khodare *et al*, 2020). Recently, attempts been made to understand the sensitivities of the SARS-CoV-2 detection real time kits in pool testing set up and feasibility. Using RealStar SARS-CoV-2 RT-PCR Kit, Lohse et al showed that they could detect a positive sample in a poll as large as 30. However, their data is limited by the fact that 30 was the largest number of pooled samples that they tested. We decided to test the efficiency of SARS-CoV2 detection in pooled samples, further increasing the number of samples in a pool using the Taqpath COVID-19 combo kit. For this, we spiked known amounts of the standard SARS-CoV-2 RNA into the pooled total RNA from healthy individuals. As per our data, our pooling techniques can detect one positive sample with RNA copies, as low as, 100 copies/ul post isolation levels in a pool of 60 samples. During the early phase of infection i.e.; within day 5 of exposure when SARS-CoV2 is vigorously replicating, the titers are upwards to 10^4^-10^6^/ul (Zou *et al*, 2020, NEJM). As per the study published in the journal NEJM by Zou *et al*; the titers in asymptomatic patients were found to be “similar” to the symptomatic patients. This suggests that pooling strategies can easily detect even the asymptomatic individuals.

Since the idea of sample pooling is to expand the screening capacity of a large population, efforts have been made to improve the process, but it still has some innate challenges, including:

1. The Ct values could be higher as compared to the individual sample Ct value, as observed by Lohse et al; this could make the correct estimation of the viral load more complicated to interpret based just based on sample dilution. These sample pooling strategies are seldom used for accurate estimation of the viral load, these typically have only binary outcomes.
2. In the absence of an internal control like RNAse P, it would be difficult to rule out the possibility that the absence of any detectable infection was due to bad sampling practices or just simply a manual error of addition. There are limited corrective measures available once the sampling is done and it reaches the lab, however, smart pooling strategies like samples overlapping between different pools could be used to minimize manual addition error. Also, with increasing use of liquid handling systems as well as AI driven sample pooling strategies aforementioned challenges could be managed well.

Given the stress on our existing testing facilities, availability of the kit and financial consideration, sample pooling strategy is a long-term solution for the businesses to reopen and function during such pandemics. This is particularly true for clinical setting which might require a frequent testing of all the staff as well as visitors.

#### Sample pooling efficiency within distributions for prevalence and positive sample allocations

Here, we constrain a max pool size of 64 samples, a maximum number of testing rounds of 3, and require individual confirmation of a positive sample.

As our objective in pooling samples (over individual testing) is to perform better (fewer) than one test per sample, we model relative^1^ reactions per sample throughout a range of prevalences:

The maximum pool size of 64 was chosen as a function of Ct for detectability. Other techniques that could improve search cost have been eliminated either due to the failure to confirm positive samples directly, or due to the total time required to identify positive samples (proxy via maximum rounds), or because of the risk introduced by human error in assembling subsequent pools.

We therefore chose a cross-pool strategy that begins with a single pool of the maximum pool size (64-pool) arranged 8×8 (wells A1:H8 on a standard 96-well microtiter plate). If the pooled sample returns positive, continues 16 parallel tests of 8 samples (8-pool) comprised of each row and each column from the 64-pool such that each sample is part of exactly two 8-pools; and finally, positive 8-pools identify the individual cells or ranges of cells of possible active samples. These are then tested individually.

The maximum number of testing rounds was chosen to trade-off a practical limitation of the time that can be expended (relative to total time/cost), to deliver test results. The effect of these constraints is that the test marginally^2^ performs better at these maximums, and thus a pool of 64 is chosen, and three rounds are prescribed for all testing where the first 64-pool result is positive. Thus, absolute reactions per positive sample is always four -- that is, all positive samples have been part of four pools: {64-all, {8-col, 8-row}, 1-individual} over three rounds.

#### Next, we demonstrate the problem of positive sample distribution within the initial pool

Best and worst-case performance of total reactions in high vs low prevalence pools of 64 samples.

In low prevalence pools (LPPs), pools <= 5% positive sample rate, nonoptimal (worst) distribution of positive samples has minimal effect on R-RPS variance.

In high prevalence pools (HPPs), those >= 12.5% positive sample rate, least optimal distribution of positive samples cost is 1.27 R-RPS (as shown in Figure-3, 81 Tests / 64 Samples), and varies 3:1 with ideal (best) distribution.

Thus, variance in reactions per sample is highly sensitive to prevalence.

We then look to identify an expected prevalence threshold that’s acceptable under worst-case performance.

#### R-RPS of Best, Worst, and Avg Positive Sample Distribution in 64-Pool

**Table 1:** Best, worst and avg reactions per sample in a pool subject to positive-sample-distribution, of a 64-sample pool.

| Prevalence | Best | Worst | AVG |
| --- | --- | --- | --- |
| 0.055 | 0.328125 | 0.515625 | 0.421875 |
| 0.060 | 0.328125 | 0.515625 | 0.421875 |
| 0.065 | 0.343750 | 0.656250 | 0.500000 |
| 0.070 | 0.343750 | 0.656250 | 0.500000 |

## Conclusions

We propose that when expected prevalence is below 6% that pooled testing may be relied upon to reduce individual testing costs by approximately half and reduce the likelihood of false positives.

With aforementioned sample pooling strategy, frequent screening of the employees can be achieved at a much lower financial burden. We based our analysis on the R-RPS, as due to its direct interpretation in financial terms. Sample pooling would not only impart a significant financial advantage but also help maximise the resource both in terms of the number of reactions needed for a large-scale screening. Given the uneven distribution of cases across US, in the area where prevalence is below 6% this strategy can be very helpful in getting the businesses started without compromising on their ability to monitor any future outbreak.

We have created a highly adaptable algorithm that is able to rapidly assist a testing lab with the protocol to be used based on the number of samples that are at the lab. This would prevent undue delays due to batching of samples and would ensure that the cost benefit aspect is not lost due to a varying number of samples that would be expected during the pandemic. It is also important to note that our algorithm is agnostic to the agent (for example: a different virus, bacteria) and will be rapidly adaptable to any new outbreak.

## Material and methods

### Human and SARS-Cov2 RNA isolation

This study was waived from the review by the Zouves Foundation Institutional Review Board (OHRP IRB00011505). Total human RNA was purified from nasal swabs from healthy human donors, tested negative for SARS-Cov2. Nasal swabs were collected using Nylon flocked nasopharyngeal swabs (Hardy Diagnostics) at our clinic by trained medical professionals. The MagMAX Viral/Pathogen II (MVP II) Nucleic Acid Isolation Kit was used to isolate total RNA as per manufacturers recommended protocol. TaqPath COVID-19 Positive Control for Taqpath RT-PCR Covid-19 Kit was used as the standard SARS-Cov2 RNA with known copy number for the assays.

### Quantitation of SARS-Cov2 RNA

Standard SARS-Cov2 RNA were spiked in the known amounts total human RNA to simulated the dilution of the SARS-Cov2 positive control sample. Quantitative real time PCR was used to estimate the SARS-Cov2 RNA from the simulated pooled samples using the Taqpath COVID-19 combo kit (Thermo Fisher Scientific). The real time PCR reactions were performed on the QuantStudio3 and Ct values for each sample was calculated using Thermo Fisher Connect Design and Analysis 2 software. Samples for which Ct values were found to be less than 40 cycle for two out of the three regions in the viral genome (S gene, N gene and ORF1ab) were considered positive in our assay.

### Data representation and analysis

The Ct values form the real time PCR experiments were plotted using Excel 2016 (Microsoft office). The number of reaction per pool in figure 2 was plotted using Matplot and Seaborn.

**Figure 1:**
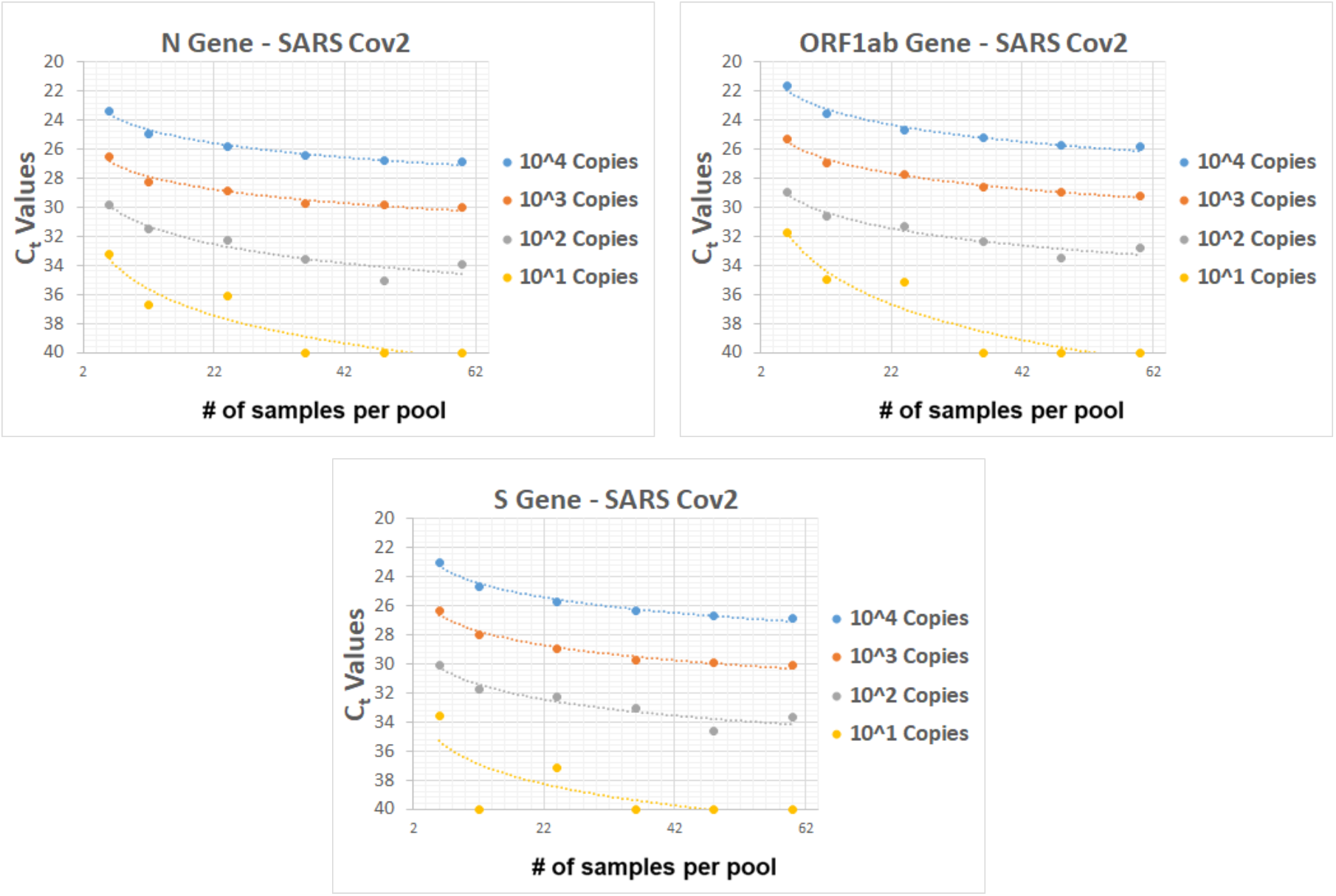
SARS-CoV2 RNA load detected in the pooled total RNA from healthy individuals, spiked with known amounts of the standard SARS-CoV-2 RNA (AcroMetrixTM Coronavirus 2019 (COVID-19) RNA Control). Ct values beyond 40 cycles were considered undetectable

**Figure 2:**
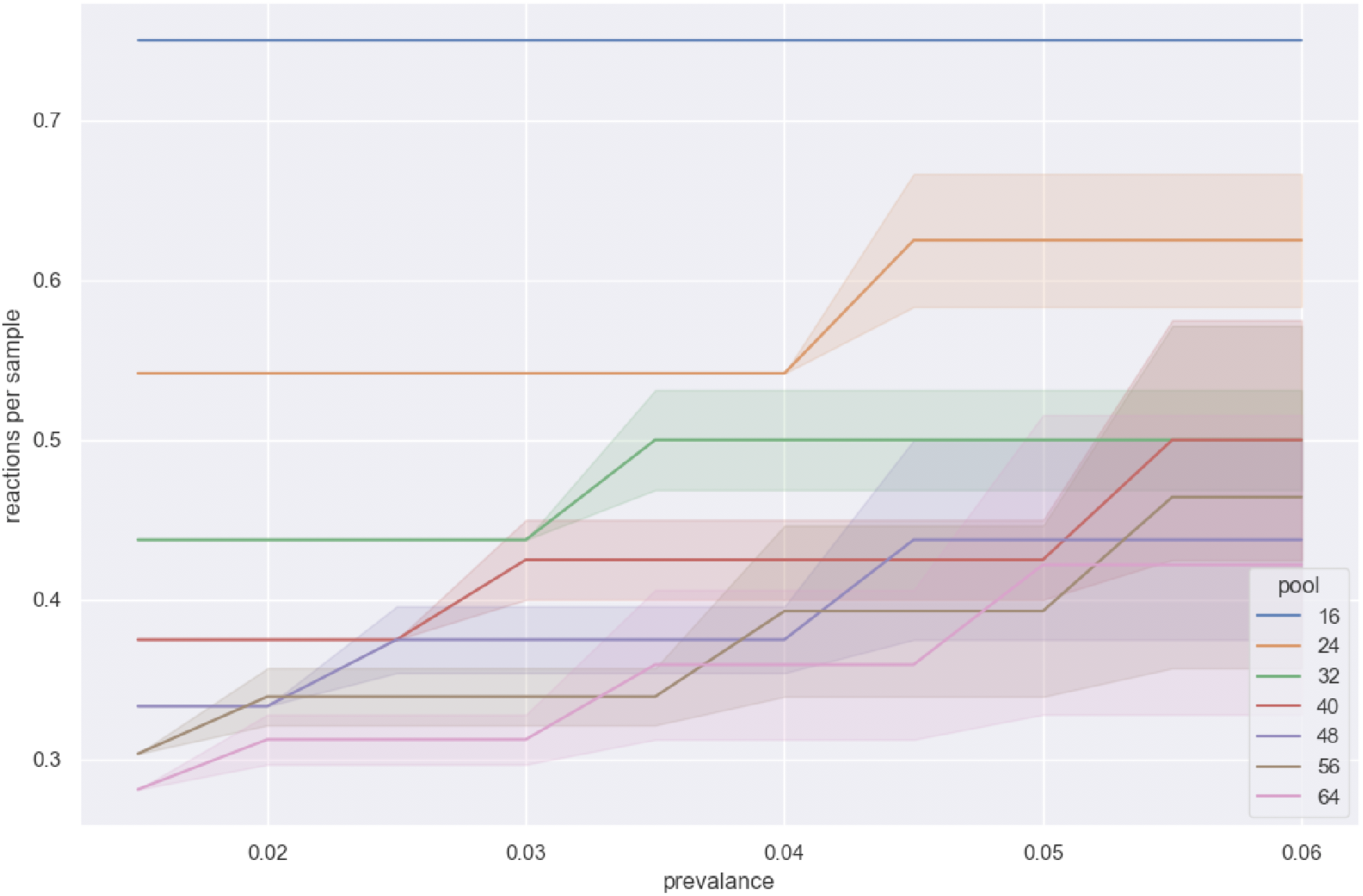
Sample pools (from 16 to 64 samples) with average test performance measured as relative reactions per sample (R-RPS), (Total Reactions / No. of Samples). Shaded region is lower and upper boundary R-RTS for best and worst case positive sample distribution (described below).

**Figure 3:**
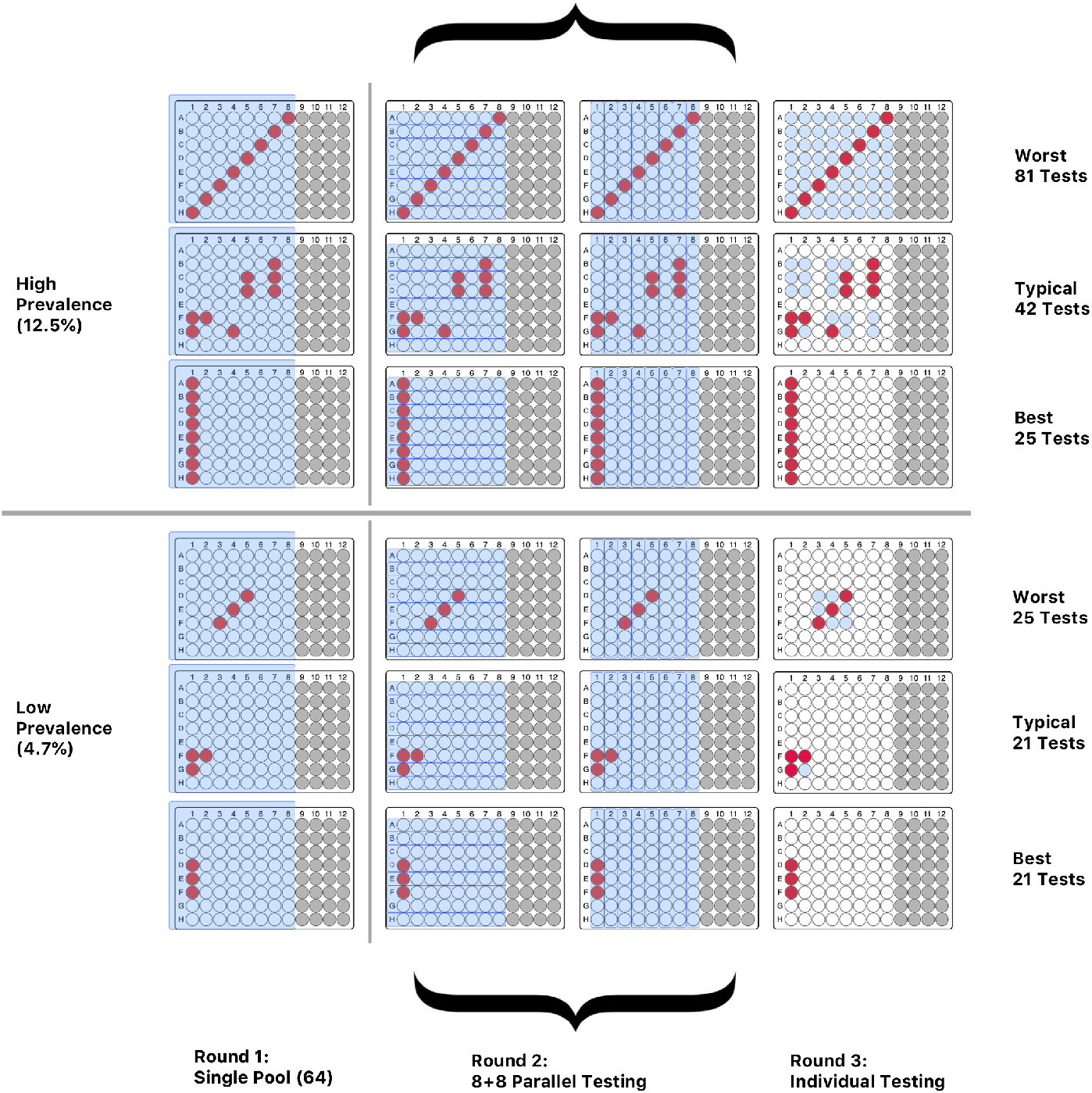
high and low prevalence comparisons are shown with three positive-sample-distributions (worst, typical (+-1SD), best), demonstrating the R-RPS variance sensitivity to prevalence.

## Data Availability

All data is submitted and available on request

## Footnotes

1 Not to be confused with absolute reactions per sample (e.g. the absolute number of times a sample has been tested, or the number of pools that a sample has been member of).

2 Overlap between pools in Figure-2 are the result of pool smaller row pool sizes (n/8).

## Notes

### Competing Interest Statement

S. Sawarkar, S. Verma, J. Sams, and S. Haran are founders at Rosa Scientific. M. Viotti, A. Victor are employed at Zouves Foundation for Reproductive Medicine

### Funding Statement

Our research was supported by funding from ZFRM and Rosa Scientific

### Author Declarations

This study was waived from the review by the Zouves Foundation Institutional Review Board (OHRP IRB00011505).

